# An epigenetic speedometer to measure Pace of Aging: FraminghamPACE

**DOI:** 10.64898/2026.07.07.26357388

**Authors:** William T Marella, Calen P Ryan, David Corcoran, Claire Eckstein Indik, Alex Furuya, Michael S Kobor, Karen Sugden, Avshalom Caspi, Terrie E Moffitt, Daniel W Belsky

**Author notes:** Correspondence to, 722 W 168^th^ St, Suite 410, New York, NY 10032.

## Abstract

Geroscience clinical trials need biomarker surrogate endpoints for healthspan. Leading candidates are omics-based composites developed from machine learning analysis of aging phenotypes including calendar age, survival, functional capacity, and Pace of Aging. Existing Pace of Aging biomarkers were developed in the Dunedin Longitudinal Study, limiting inference about the strengths and weaknesses of the method as distinct from the Study, a unique single-year birth cohort followed through midlife with near-perfect retention and uniform measurement of multiorgan-system function across two decades of follow-up. We adapted our Pace of Aging method for mixed-age cohorts with variable follow-up of organ-function measures and applied it to develop a novel DNA methylation biomarker of Pace of Aging in data from the Framingham Heart Study Offspring Cohort, FraminghamPACE. Validation analyses across four independent cohorts and one clinical trial establish advantages for the Pace of Aging method in developing biomarkers that are both predictive of healthspan and responsive to geroprotective intervention.

## INTRODUCTION

The global population is aging, threatening decreased productive capacity due to shrinking workforces and steep increases in the costs of health and social care. The chronic, disabling conditions of old age develop over many years, beginning well before clinical diagnosis. Therefore, preventative intervention must also begin early. However, longitudinal follow-up from early intervention to disease/disability onset is impractical. Drug developers cannot afford to wait decades to learn whether new treatments work. Physicians and patients seeking personalized care require feedback over the course of months, not decades. This gap has driven interest in biomarkers of aging that can function as surrogates for healthspan in clinical trials and guide patient care.

The leading biomarkers of aging are composite variables that integrate information from dozens to hundreds of variables, often derived from multiplex “omics” assays^1,2^. Data integration in these biomarkers is achieved using machine learning methods that fit a model using high-dimensional datasets of biological features, such as the output of an omics microarray, to predict an outcome that represents aging. The result is an algorithm that can be applied to new datasets, such as baseline and follow-up measurements in a clinical trial.

The outcome most often used to develop aging biomarkers is age itself, i.e. years lived since birth^3,4^. An alternative approach requiring additional data is to set the outcome as survival time, i.e. years of remaining life^5^. These approaches have intuitive appeal. However, in humans, relying on years lived as an outcome introduces a range of biases^6^, most prominently confounding of aging with survival; humans in their 70s and beyond are, by definition, successful agers, having outlived most of their peers. The results of machine learning analysis differentiating older from younger people could therefore reflect not only aging-related biological damage, but also resilience. Using survival time as an outcome together with statistical controls for variation in age at measurement of biological variables helps to resolve this limitation and some others; aging biomarkers developed from machine learning analysis using survival time as the outcome are among the best validated in the literature^7^. Nevertheless, the focus of these biomarkers on lifetime accumulation of risk has the potential to limit their sensitivity to interventions that slow aging, but do not reverse it.

An alternative approach that may overcome this limitation is to apply machine learning to an outcome that represents something closer to what many interventions aim to modify: the current rate of aging-related biological deterioration. We developed such a measure, Pace of Aging, by modeling changes over 20 years of follow-up in a panel of organ-function measurements among participants in the Dunedin Longitudinal Study, a single-year birth cohort followed through midlife with near-perfect retention^8,9^. Then, following methods established in studies to develop time-since-birth and survival-time biomarkers from blood DNA methylation^4,5,10^, we used Pace of Aging to develop the DNA methylation biomarker DunedinPACE^11^. This new DNA methylation biomarker of Pace of Aging performed similarly to the best of the survival-time biomarkers in prediction of incident disease, disability, and mortality^11^. Critically, it also proved more sensitive to the effects of calorie restriction^12^, the intervention best established to slow aging in a range of laboratory models^13^.

If using Pace of Aging in machine learning analysis to develop aging biomarkers can yield more sensitive endpoints for clinical trials, this would be consequential for the field. However, there are alternative explanations for the calorie restriction trial result. The participants in the trial (CALERIE^12^) were healthy midlife adults, similar to the Dunedin Study members whose data were used to develop DunedinPACE. In contrast, the leading survival-time biomarker, the GrimAge epigenetic clock^10^, was developed using data from older adults, many of whom had prevalent chronic disease. The critical factor could therefore be similarities between the participants whose data were used to develop the biomarker and the participants in the clinical trial. Consistent with this alternative hypothesis, two subsequent trials found that nutritional-supplement interventions slowed age-dependent increases in both Pace of Aging and survival-time biomarkers in older adults^14,15^.

To adjudicate between these competing hypotheses, biomarker design vs. demographic similarity, we developed a novel Pace of Aging biomarker in the same older-adult cohort used to develop GrimAge and tested its response to intervention in the CALERIE trial. We obtained data from the Framingham Heart Study Offspring Cohort^16^. We used measurements taken over the first three decades of follow-up in that study to devise a new Pace of Aging measurement. We then modeled this new Pace of Aging from DNA methylation data collected at the end of the follow-up period, the same DNA methylation data used to develop GrimAge. We applied the resulting model to data from four observational studies to establish technical reliability, patterns of change with aging, and prediction of disease, disability, and mortality, and to the CALERIE trial to test responsiveness to calorie restriction intervention.

## RESULTS

### Pace of Aging in the Framingham Heart Study ODspring Cohort

The key barrier to development of Pace of Aging biomarkers is the requirement for longitudinal data. The method requires observations of multiple organ systems at three or more time points over follow-up intervals of sufficient length to observe aging-related decline^8^. The Dunedin Study Pace of Aging analysis was made possible by the study’s birth-cohort design (all participants are the same age) and its harmonized collection of 19 blood-chemistry and organ-function tests from the complete cohort across four assessment waves spanning two decades^8,9^. We previously introduced methods to account for variation in participant ages and changes in measurement protocols^17^. To measure Pace of Aging in the Framingham Heart Study Offspring Cohort, we further adapted our method to accommodate biomarker data collected on variable schedules (i.e. different time points for different biomarkers).

Measurements used to define Pace of Aging were collected across the first 8 waves of follow-up spanning three decades (1971-2008). We included measurements taken at least twice on >500 men and >500 women. Lipids and blood pressure were excluded due to initiation of medical management during follow-up in many participants. We curated data on 17 blood-chemistry and organ function tests reflecting vascular, inflammatory, renal, metabolic, pulmonary, hepatic, and hematologic functioning (**Figure 1; Supplemental Table 1**). Data were processed for analysis following our established procedures^8,17^. Briefly, we applied log transformation to reduce skew, scaled values to a Z distribution, and reversed values for measurements known to decline with age. Consistent with prior Pace of Aging analyses, participants exhibited a rising burden of aging-related risk across organ systems over follow-up (**Figure 1**).

**Figure 1:**
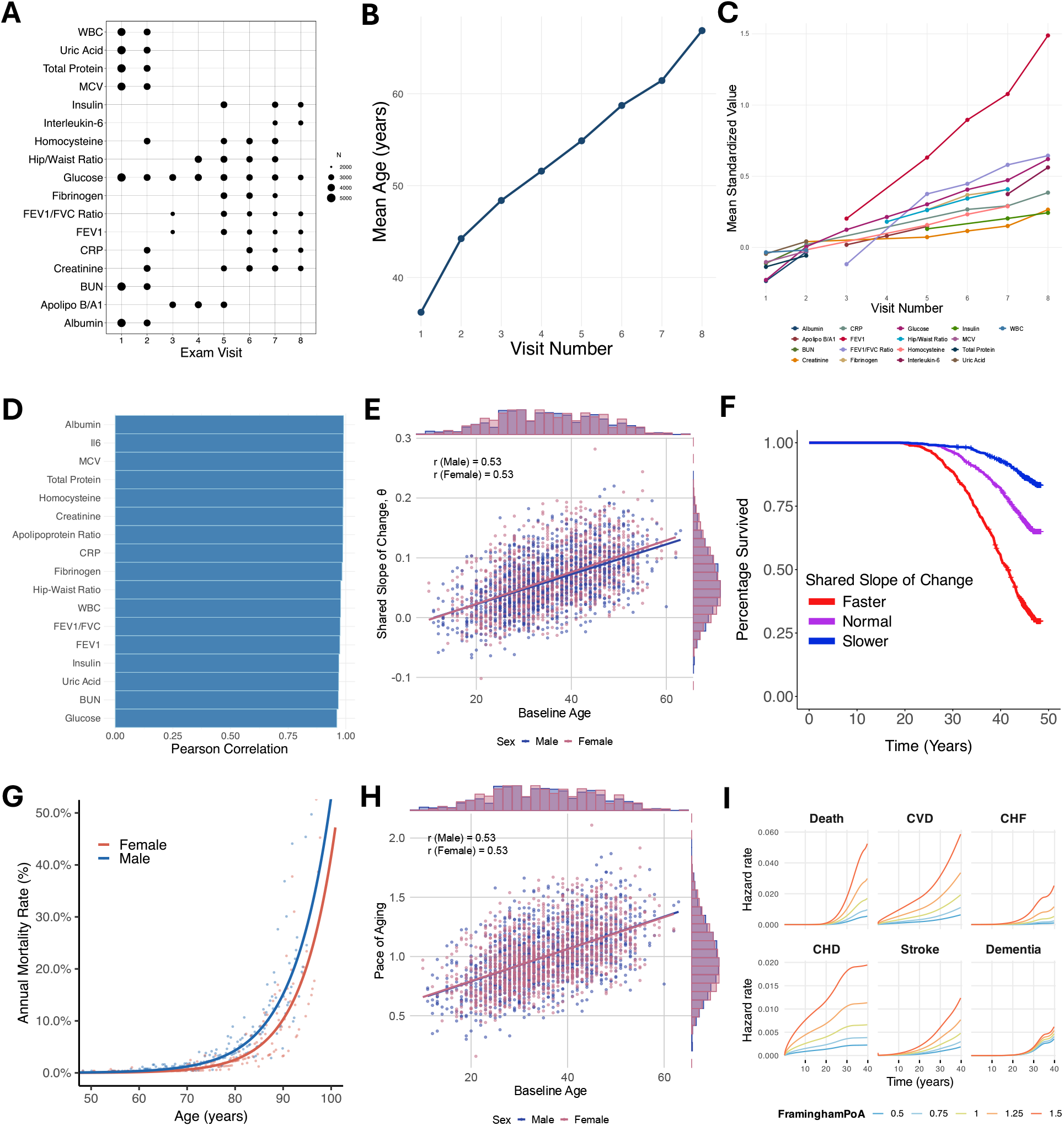
Measurement of Pace of Aging in the Framingham Heart Study ODspring Cohort. The figure illustrates analyses to measure Pace of Aging in the Framingham Heart Study Offspring Cohort. **(A)** Number of biomarker measurements across the eight FHS Offspring exam visits for the 17 biomarkers considered in the Pace of Aging model. Point size reflects the number of participants with available measurements at each visit. **(B)** Mean participant age at each of the eight exam visits. **(C)** Mean standardized trajectories of each of the 17 biomarkers across exam visits. **(D)** Leave-one-biomarker-out sensitivity analysis. Each bar shows the Pearson correlation between the full 17-biomarker shared-slope-factor and shared-slope factors obtained after refitting the hierarchical linear model with the indicated biomarker removed. All correlations exceeded 0.96, indicating that no single biomarker had substantial impact on the model estimate of the shared slope. **(E)** Shared slope of change (θ) from the hierarchical model plotted against baseline age, stratified by sex. θ shows moderate age patterning (r = 0.53 in both males and females). **(F)** Kaplan-Meier survival curves stratified by θ groups defined as faster (> mean + 0.5 SD), normal (within ±0.5 SD of the mean), and slower (< mean − 0.5 SD), demonstrating that θ captures mortality-relevant variation in aging rate. **(G)** Sex-specific Gompertz mortality models fit to the FHS participants with assigned Pace of Aging values. Smoothed curves show the fitted Gompertz functions; points show empirical annual mortality rates computed in 0.2-year age bins. **(H)** Framingham Pace of Aging plotted against baseline age by sex. Framingham Pace of Aging is a rescaled transformation of θ and retains the same age patterning (r = 0.53 in both sexes), centered so that a value of 1 corresponds to the population-typical pace. **(I)** Associations of Framingham Pace of Aging (FraminghamPoA) with six health outcomes in FHS: all-cause mortality, cardiovascular disease (CVD), congestive heart failure (CHF), coronary heart disease (CHD), stroke, and dementia. Curves illustrate hazard functions for each outcome for Pace of Aging values of 0.50, 0.75, 1.00, 1.25, and 1.50. Hazard functions were estimated using the Royston-Parmar spline method with three internal knots and covariates for age and sex. Curves show predicted hazard rates over follow-up time. Higher Framingham Pace of Aging was associated with elevated hazards across all six outcomes.

Pace of Aging analysis proceeded in three steps. First, we fit a hierarchical model^18^ to the longitudinal blood-chemistry and organ-function data (n=4,988; **Supplemental Table 2**). This approach allowed us to model all the data simultaneously and to estimate a factor representing the shared slope of change across organ systems for each participant (**Methods**). To ensure the resulting factor did not depend on any individual organ system, we conducted a leave-one-out analysis. We re-estimated the hierarchical model omitting each measurement in turn and correlated the resulting factors with the original factor estimated from the complete data. The factors resulting from the leave-one-out analysis were very similar to the original factor (Pearson r>0.96; **Figure 1**), suggesting that no single organ system had disproportionate influence.

Second, we scaled factor scores for men and women by the corresponding sex-specific mean in the cohort to compute Pace of Aging ratios (years of biological decline per calendar year; **Figure 1**). Finally, we re-scaled the ratios so that the implied years of biological change across the ∼30y follow-up (i.e. 30 for Pace of Aging=1, 60 for Pace of Aging=2) was consistent with the sex-specific survival function in the cohort (**Figure 1**). The scaled Pace of Aging value was normally distributed around 1, correlated with age, and showed a strong association with incidence of chronic disease and mortality over follow-up (**Figure 1, Supplemental Table 3**).

### FraminghamPACE

We next modeled our Pace of Aging phenotype from DNA methylation data collected at the end of the follow-up period to create a new epigenetic speedometer of aging, FraminghamPACE (with PACE standing for Pace of Aging Computed from the Epigenome). Analysis included participants with Pace of Aging and DNA methylation data (n=2,069; **Supplemental Table 2**). Briefly, we used ensemble regularized regression^19,20^ to fit Pace of Aging to a set of CpG sites selected to ensure adequate consistency of measurement across generations of Illumina DNAm arrays (**Supplemental Table 4**; see **Methods** for details). We fit two models. The first model used the conventional approach for developing epigenetic clocks, elastic-net regression^19^. This approach selects a subset of the total CpG sites considered in the analysis to form the final algorithm. The second model used an alternative approach motivated by prior studies suggesting algorithms that distribute weights across a larger number of CpG sites have superior technical reliability^21^. This alternative approach, ridge regression^22^, retains the entire set of CpG sites included in the analysis in the final algorithm. Hereafter we refer to these alternative versions of FraminghamPACE as “ElasticNet” and “Ridge”.

The FraminghamPACE epigenetic speedometers explained slightly less than half the variance in our Pace of Aging phenotype (ElasticNet hold-out R^2^=0.47; Ridge hold-out R^2^=0.46; **Supplemental Table 5**). The ElasticNet model included 1,123 CpG sites. The Ridge model included 14,465 CpG sites (all sites included in analysis). Contributions of individual CpG sites to the speedometers are illustrated in **Figure 2**. Despite moderate fits, both FraminghamPACE versions were comparable or better predictors of healthspan and lifespan in Framingham participants relative to the original Pace of Aging phenotype (**Figure 2, Supplemental Table 3)**. This result is similar to what we observed for DunedinPACE in the Dunedin Study cohort^11^. The consistency of the increment in effect sizes between the DNA methylation biomarkers and the original Pace of Aging suggests that FraminghamPACE is successfully distilling biological-aging signal and screening out noise due to measurement error.

**Figure 2:**
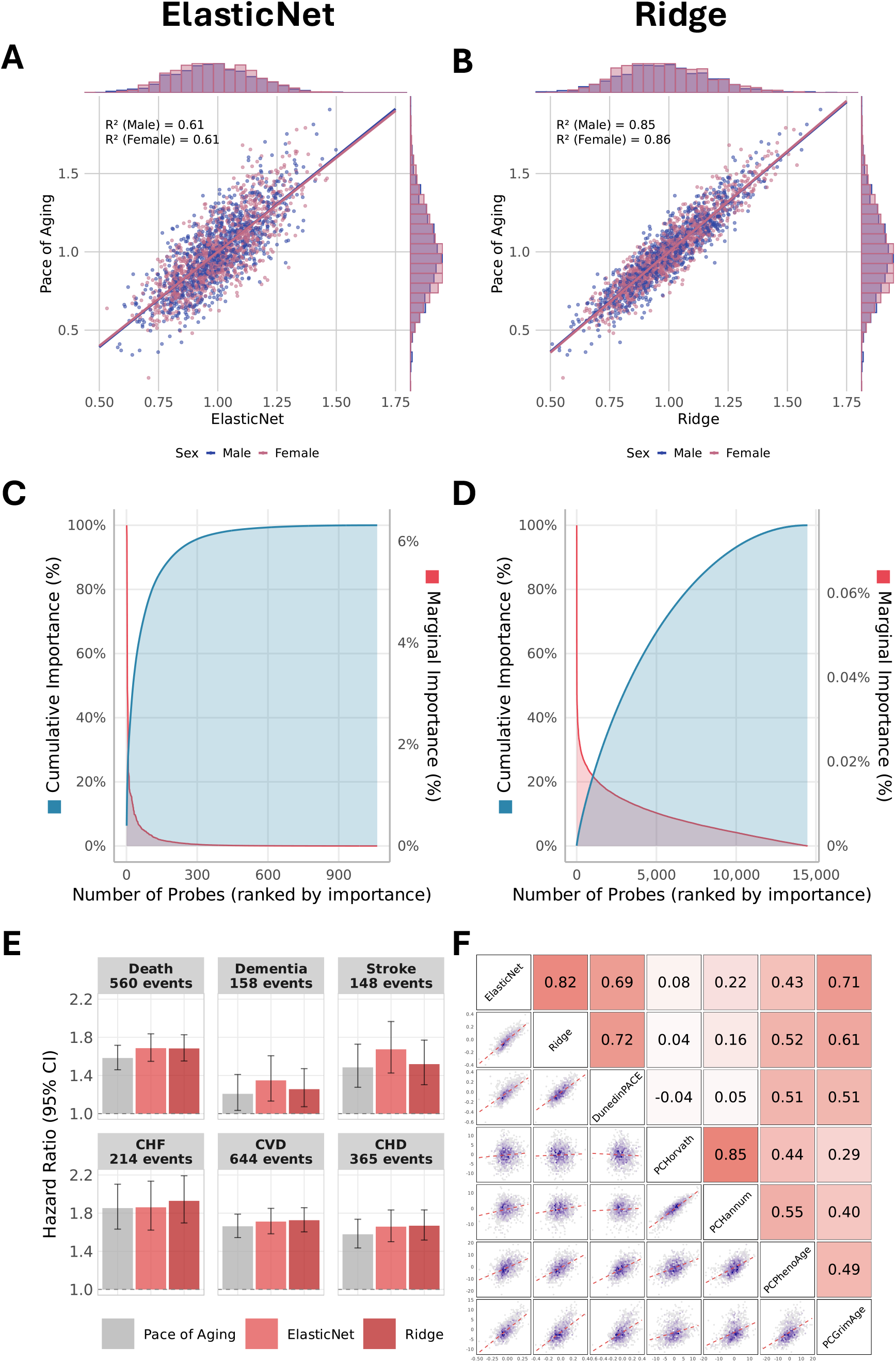
Development of FraminghamPACE. The figure illustrates analyses to develop the FraminghamPACE ElasticNet and Ridge epigenetic speedometers. **(A)** In-sample fit of FraminghamPACE ElasticNet against the underlying Framingham Pace of Aging values. Points show individual participants colored by sex; dashed lines show sex-specific regression fits (R^2^ = 0.61 in both sexes). **(B)** Equivalent in-sample fit for FraminghamPACE Ridge (R^2^ = 0.85 in males, R^2^ = 0.86 in females). **(C)** Probe importance distribution for FraminghamPACE ElasticNet. CpG probes are ranked by importance, defined as the absolute model coefficient weighted by the probe’s empirical standard deviation and normalized to sum to 1. The blue curve shows cumulative importance (left axis) and the red curve shows marginal importance per probe (right axis). The elastic net solution is sparse, with a small number of probes accounting for the majority of predictive signal. **(D)** Equivalent probe importance distribution for FraminghamPACE Ridge. The ridge solution distributes importance across many more probes. **(E)** Hazard ratios per 1-SD increment (95% CI) from Cox models for six time-to-event outcomes in FHS — all-cause mortality, dementia, stroke, CHF, CVD, and CHD — comparing the underlying FraminghamPoA (gray) to FraminghamPACE ElasticNet and FraminghamPACE Ridge (red). The DNAm proxies recover or exceed the prognostic performance of the raw Pace of Aging measure across outcomes. **(F)** Correlations among FraminghamPACE ElasticNet, FraminghamPACE Ridge, DunedinPACE, PCHorvath, PCHannum, PCPhenoAge, PCGrimAge in CARDIA. Upper triangle shows Pearson correlations; lower triangle shows corresponding binned scatterplots.

We next evaluated the resulting FraminghamPACE speedometers in a series of external validation datasets. We first confirmed technical reliability using datasets of replicate DNA samples from the LOLIPOP and CARDIA cohorts. We then conducted analysis of correlation with existing epigenetic clocks and changes with aging in longitudinal data from the CARDIA cohort. We tested prediction of healthspan and lifespan in the HRS and WHI cohorts. Finally, we evaluated responsiveness to intervention in the CALERIE Trial. Participant characteristics in these datasets are reported in **Supplemental Table 6**.

### FraminghamPACE has excellent technical reliability

Reliability of FraminghamPACE was determined using intraclass correlations (ICCs) for n=36 replicates from the London Life Sciences Prospective Population (LOLIPOP) study (GSE55763^23,24^) and n=38 replicates from the Coronary Artery Risk Development in Young Adults (CARDIA) (**Methods**; **Supplemental Methods**). FraminghamPACE showed excellent technical reliability in replicates from LOLIPOP (ICCs ≥0.97) and CARDIA (ICCs ≥0.95). Full results and comparison with other DNAm biomarkers of aging are shown in **Supplemental Figure 1** and **Supplemental Table 7**.

### FraminghamPACE is correlated with other DNAm biomarkers of aging

To evaluate similarity between the new FraminghamPACE speedometers and published epigenetic clocks, we analyzed data from the CARDIA cohort collected at the time of the year-15 follow-up, when participants were in their mid-30s to mid-40s (N=2,023). FraminghamPACE values showed strong correlations with DunedinPACE (r∼0.7 for both ElasticNet and Ridge) and with age-residualized values of the PCGrimAge clock (r=0.7 for ElasticNet, r=0.6 for Ridge). Correlations were weaker for age-residuals of the PCPhenoAge clock (r=0.4-0.5) and the Horvath and Hannum clocks (r=0.04–0.22). Correlations and scatterplots illustrating associations are graphed in **Figure 2**.

### FraminghamPACE indicates that the aging process accelerates across young adulthood through midlife

As measured by survival probabilities, human aging accelerates as we grow older; after the age of 30, mortality risk doubles every ∼8y for humans. However, most biological age models assume linear change^25^. Our Pace of Aging metric already reflects linear change. An increase with advancing calendar age therefore implies acceleration in Pace of Aging. In the Framingham cohort, older participants had faster Pace of Aging as compared to younger participants (r=0.53; **Figure 1**). We tested aging-related acceleration of FraminghamPACE in n=2,023 participants with longitudinal DNAm data in the CARDIA cohort (four time points of measurement over 15y of follow-up). Participants’ FraminghamPACE values increased as they grew older (for ElasticNet, per-year b=0.002 95% CI [0.0018-0.0022]; p<0.001; for Ridge, per-year b=0.003 [0.0027-0.0031]; p<0.001; **Figure 3, Supplemental Table 8**), similar to previous observations for DunedinPACE^11^ and consistent with the hypothesis that aging accelerates across adulthood.

**Figure 3:**
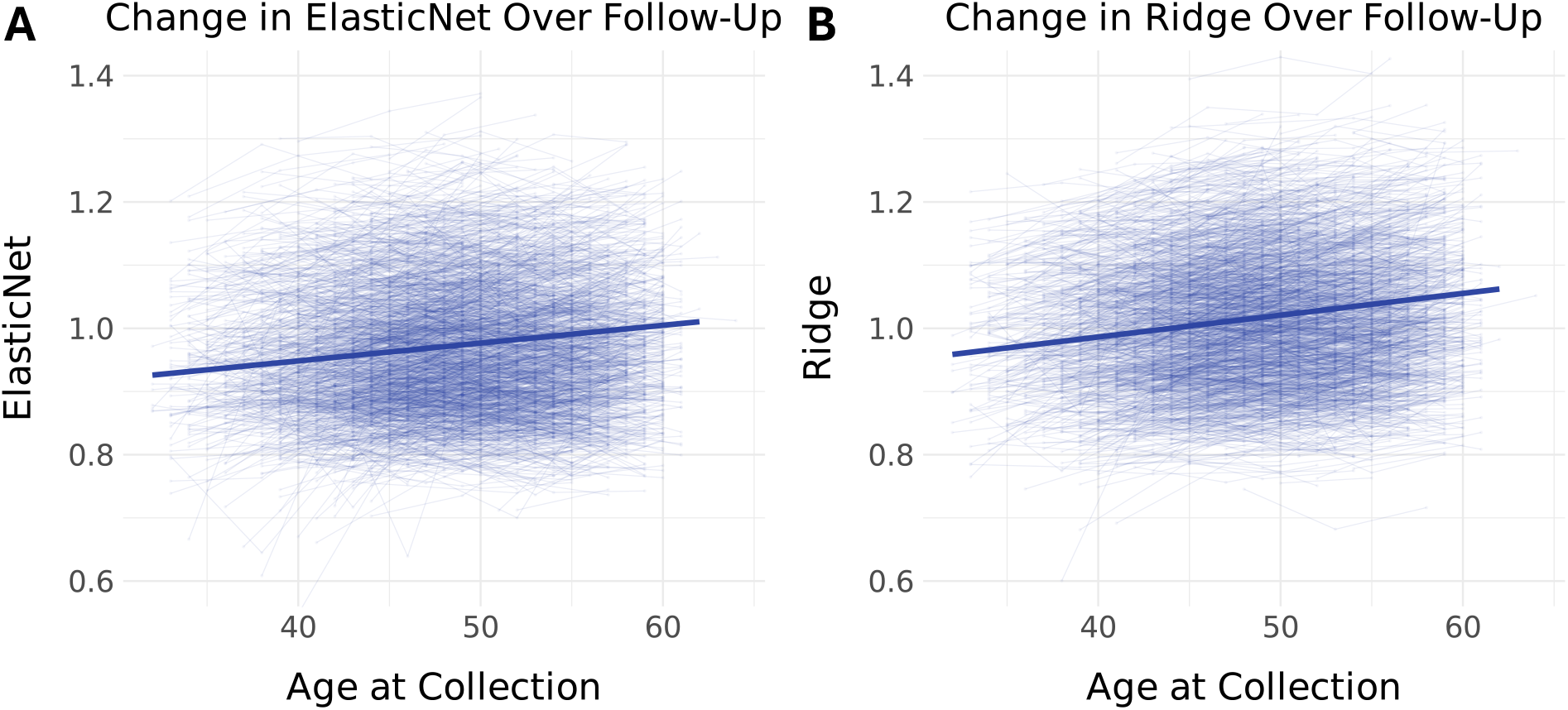
Repeated measures analysis of FraminghamPACE indicates accelerating Pace of Aging across adulthood. The figure shows repeated-measures DNA methylation data from the Coronary Artery Risk Development in Young Adults cohort (CARDIA, n=2,023). Participants provided DNA samples at up to four time points spanning the year 15-30 follow-up visits. We computed FraminghamPACE from the DNA methylation data and plotted each participant’s personal trajectory in a “spaghetti”-style plot, superimposing the cohort mean trend as estimated from within-subjects regression. Data for the ElasticNet model are plotted in **Panel A**. Data for the Ridge model are plotted in **Panel B**. The figure shows participants’ Pace of Aging values increased as they grew older.

### Adults with faster FraminghamPACE have shorter healthspan and lifespan

To establish predictive validity for FraminghamPACE, we conducted analysis of survival and incident chronic disease and disability in data from the US Health and Retirement Study (HRS, n=3,483, 6y of follow-up) and Women’s Health Initiative (WHI, n=1,835, 25y of follow-up), with outcomes detailed in **Supplemental Table 9**. Effect sizes for FraminghamPACE were similar to effect sizes for DunedinPACE and PCGrimAge (**Figure 4, Supplemental Table 10**). Results were similar after covariate adjustment for cellular composition of DNA samples, smoking history, and body mass index (**Supplemental Table 11**).

**Figure 4:**
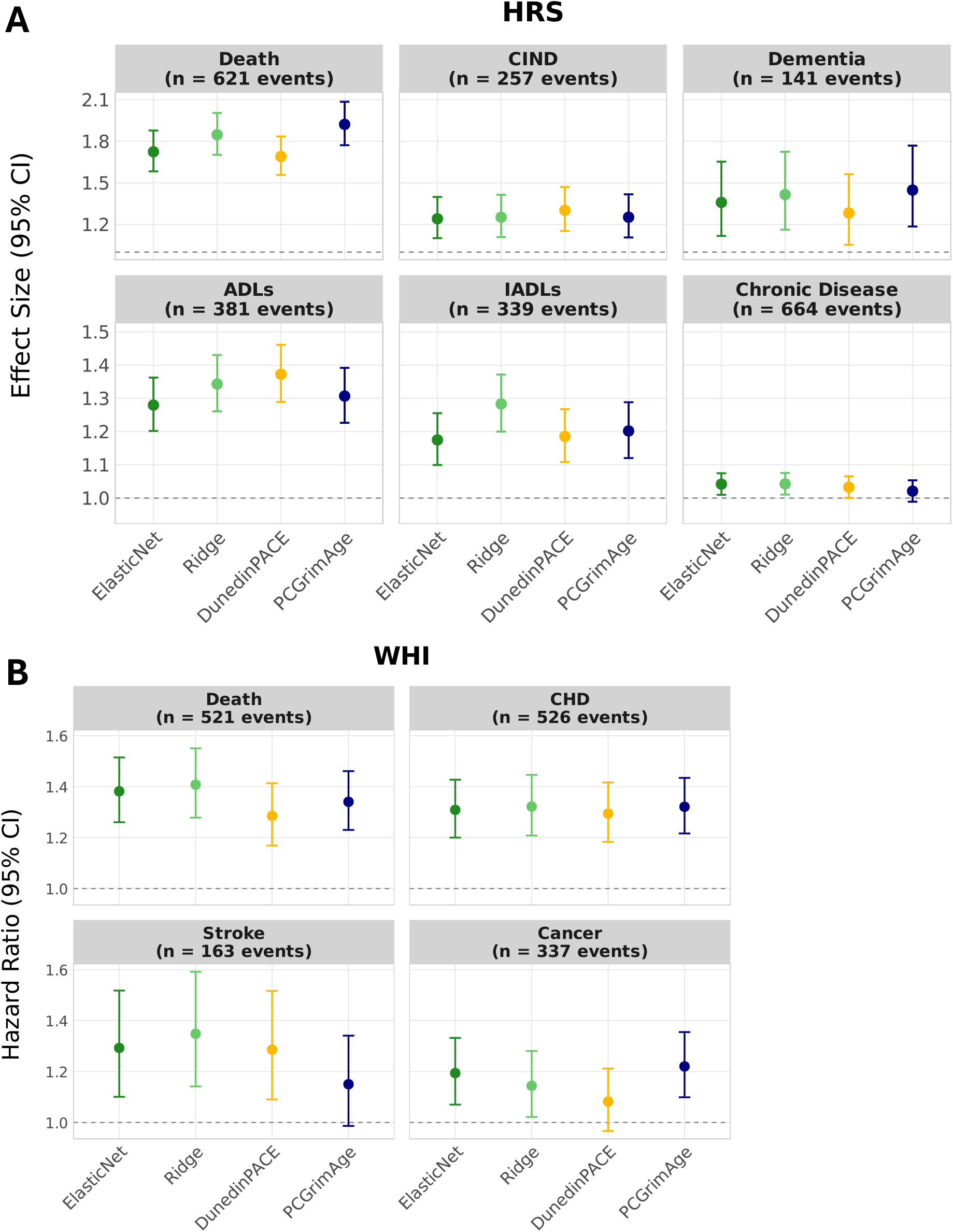
Analysis of healthspan and lifespan in the US Health and Retirement Study (HRS) and Women’s Health Initiative (WHI) cohorts. **Panel A** plots effect sizes for incident mortality, cognitive impairment without dementia (CIND), dementia, limitations to activities of daily living (ADL), limitations to instrumental activities of daily living (IADL), and chronic disease in the HRS cohort (n=3,483, up to 6y of follow-up). Effect sizes are hazard ratios for mortality, risk ratios for CIND and dementia, and incidence rate ratios for ADLs, IADLs, and chronic disease counts. The number of ‘events’ reports the number of observed outcome events for mortality, the number of participants classified as CIND or dementia at follow-up for cognitive outcomes, and the number of participants with a nonzero follow-up count for ADL limitations, IADL limitations, or chronic disease. **Panel B** plots hazard ratios for incident mortality, coronary heart disease (CHD), stroke, and cancer in the WHI cohort (n=1,835, up to 25y of follow-up). Effect sizes are also plotted for DunedinPACE and PCGrimAge for comparison purposes.

### CALERIE Trial Analysis of FraminghamPACE

CALERIE randomized N=220 healthy, non-obese adults to 2y of regular diet (“ad libitum”, AL) or 25% calorie restriction (CR)^26^. DNA methylation data were derived from blood samples collected at baseline and 12- and 24-month follow-ups. Analysis included 128 CR and 69 AL participants who provided DNA at baseline and one or more of the follow-up assessments. We tested the impact of CALERIE intervention on participants’ FraminghamPACE value using the same intent-to-treat analysis as in our original investigation^12^.

Similar to DunedinPACE and distinct from PCGrimAge and other clocks, participants in the CR intervention group experienced slowed FraminghamPACE compared with those in the AL control group (for ElasticNet, 12mo follow-up b=-0.23 95% CI [-0.41, -0.06], p=0.010, 24mo follow-up b=- 0.34 [-0.52, -0.16], p<0.001; for Ridge, 12mo follow-up b=-0.17 [-0.28, -0.05], p=0.005, 24mo follow-up b=-0.11 [-0.23, 0.01], p=0.069; **Figure 5, Supplemental Table 12**). For the ElasticNet model, treatment effects were somewhat smaller at 12 months and somewhat larger at 24 months relative to DunedinPACE. For the Ridge model, treatment effects were somewhat smaller relative to DunedinPACE at both time points. Results were similar in models that added covariate adjustment for DNAm-estimated cell composition of blood samples (**Supplemental Table 13**).

**Figure 5:**
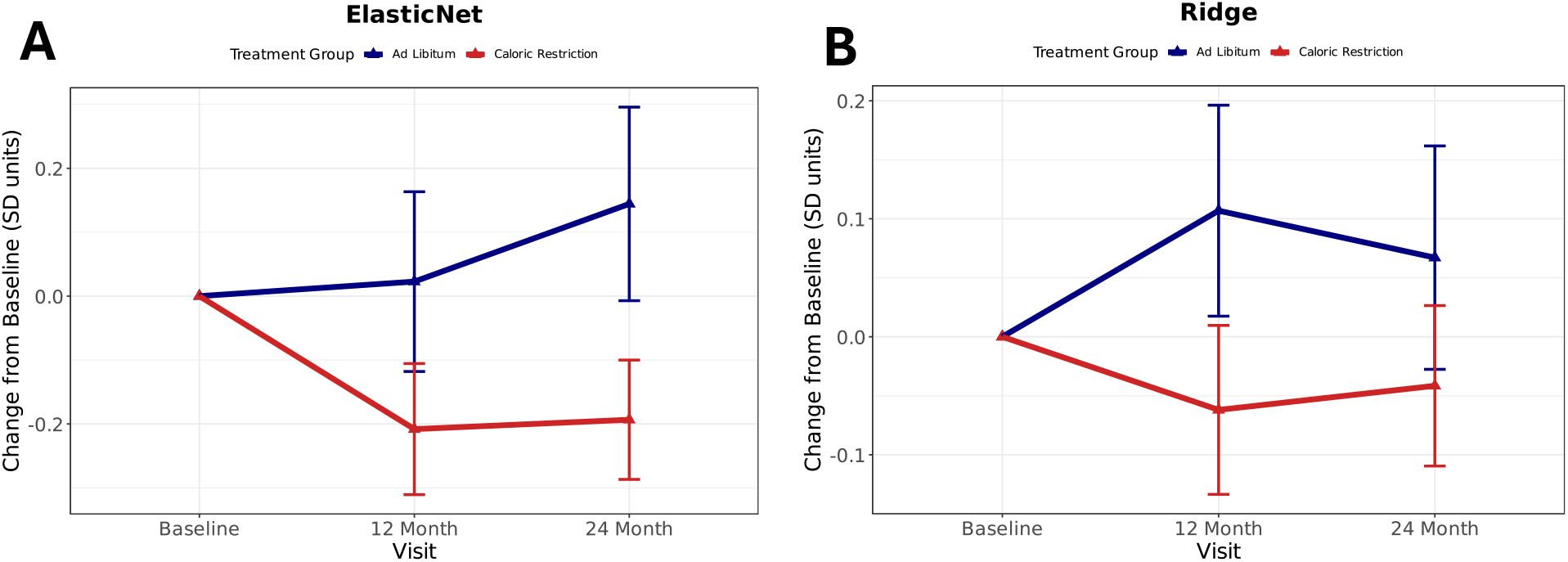
CALERIE trial analysis of FraminghamPACE. The figure plots change scores and 95% confidence intervals for FraminghamPACE among CALERIE trial participants in the ad libitum control group (blue, n=69) and calorie restriction treatment group (red, n=128). Change scores reflect differences from baseline scaled in standard-deviation units. **Panel A** shows results for the ElasticNet model. **Panel B** shows results for the Ridge model.

### DISCUSSION

We conducted analysis to develop a novel DNA methylation (DNAm) biomarker of the Pace of Aging in the Framingham Heart Study Offspring Cohort. When applied in independent cohorts, this novel biomarker (1) demonstrated exceptional technical reliability; (2) revealed a pattern of accelerating Pace of Aging with advancing age, replicating a finding first observed for our original Pace of Aging biomarkers developed in the Dunedin Study^11,27^; and (3) performed similarly to leading DNA methylation biomarkers of aging in analyses predicting healthspan and lifespan in the US Health and Retirement Study and the Women’s Health Initiative. In analysis of a randomized controlled trial of calorie restriction in healthy, non-obese humans (CALERIE), our novel Pace of Aging biomarker was slowed by calorie restriction, parallel to our original Pace of Aging biomarker and distinct from the null response of DNAm biomarkers developed to predict age and survival time, including the GrimAge epigenetic clock, developed from the same DNAm data we used in our study^12^. Together these findings establish a novel DNAm biomarker of the Pace of Aging for application in population and translational geroscience.

Our work has practical and theoretical implications. On a practical level, the novel Pace of Aging biomarker introduced here, FraminghamPACE, provides researchers with a means to replicate evidence of Pace of Aging signal generated from analysis of DunedinPACE using a parallel biomarker developed in an independent dataset. Epigenetic clocks and other omics-based biomarkers remain challenging constructs to interpret. Beyond the “first generation” of epigenetic clocks developed to predict calendar age, there are few examples of the same method being used to develop clocks from different populations or data. This limits inference about the constructs the clocks are designed to measure and also our ability to understand how properties of the data used to develop a clock relate to its performance in different settings. By introducing a second Pace of Aging epigenetic clock developed from an older population and using a different set of clinical parameters, we provide for more robust evaluation of hypotheses about how exposures or interventions may affect Pace of Aging.

With respect to theory, our results support the validity of Pace of Aging as a biomarker construct in geroscience^6,8,28^. FraminghamPACE was developed from DNA collected much later in the life course as compared with DunedinPACE; on average FHS participants were 67 years old at the time of DNA collection, as compared with 45 in the Dunedin Study. The set of clinical measurements used to model Pace of Aging was also substantially different; of the 17 clinical measurements analyzed in the Framingham cohort, only 7 overlapped with the 19 included in the Dunedin-Study Pace of Aging. Despite these differences, FraminghamPACE was highly correlated with DunedinPACE and performed similarly in analysis predicting healthspan and lifespan as well as in analysis testing response to calorie restriction. These results suggest that the DunedinPACE and FraminghamPACE epigenetic clocks are measuring a common biological construct that generalizes beyond any specific group of research participants or battery of clinical tests. Efforts are needed to expand Pace of Aging measurement across additional cohorts with the goal of increasing sample size for modeling epigenetic and other omics-based biomarkers as well as conducting analyses to understand underlying biology. The data and methods reported here provide a roadmap for this effort.

The specific result in CALERIE resolves an outstanding question about the relative performance of different epigenetic clocks in that trial. Our original finding^12^ could not exclude the possibility that the Pace of Aging biomarker developed in the Dunedin Study was more sensitive to CALERIE intervention as compared with other epigenetic clocks because of similarities between participants in the Dunedin Study and those in the trial. At the time of Pace of Aging measurement, Dunedin Study participants were midlife adults mostly free of chronic disease, similar to the inclusion criteria for CALERIE. In contrast, GrimAge and PhenoAge were developed from cohorts of older adults, many of whom had comorbid chronic conditions. Here, we found that a Pace of Aging epigenetic clock developed in the same Framingham Heart Study cohort used to develop GrimAge responded to CALERIE intervention in parallel to the original Dunedin Study Pace of Aging epigenetic clock. This result rules out participant characteristics as the explanation for the greater sensitivity of the Dunedin Study clock. Instead, it supports the interpretation that the Pace of Aging method may yield epigenetic clock biomarkers with greater sensitivity to certain interventions.

Subsequent to our original CALERIE analysis, epigenetic clock analyses of two further randomized trials testing interventions to promote healthspan, DO-HEALTH^14^ and COSMOS^15^, found evidence that, in addition to DunedinPACE, GrimAge and PhenoAge were also responsive to intervention. DO-HEALTH tested effects of omega-3 and vitamin D supplementation along with exercise in 777 healthy older adults over 3 years (positive findings were for omega-3 supplementation and, in the case of PhenoAge, for a combination of interventions). COSMOS tested effects of multivitamin and cocoa extract supplementation in 958 healthy older adults over 2 years (positive findings were for multivitamin supplementation). Whether the sensitivity of PhenoAge and GrimAge in these trials owes to participant characteristics or features of the interventions is unknown. Moreover, in the case of all three trials, at the time of epigenetic clock analysis, data were not available on long-term health outcomes such as chronic disease incidence. Therefore, it remains unknown whether epigenetic clock responses to intervention reflect accurate forecasts about future healthspan^29^.

Beyond the CALERIE result, findings here advance methods for Pace of Aging measurement and Pace of Aging biomarker development. Regarding methods for measurement of Pace of Aging, the hierarchical modeling approach introduced here can expand the range of cohorts in which measurement is possible. Pace of Aging analysis requires longitudinal data in which most or all biomarkers are measured at three or more time points. Our original method modeled rates of change in each constituent biomarker independently. A consequence is that data integration is challenging in cases where biomarkers are measured over different sets of time points, as is common in many long-running longitudinal studies and electronic health record databases. By modeling each biomarker as a random effect and estimating a shared slope factor across the set of biomarkers, the hierarchical model makes possible integration of biomarker data collected across variable sets of time points. This modeling approach also brings the method into closer alignment with geroscience theory positing a latent set of cellular-level aging processes driving change across multiple organ systems^30,31^.

Regarding epigenetic clock development, we introduced three innovations extending our previous work. First, we updated our curation of CpGs to ensure the clock can be computed using current generations of Illumina microarrays. In our original Pace of Aging epigenetic clock, we curated CpG sites demonstrating relatively higher technical reliability across the Illumina 450k and EPICv1 arrays^11,32^. Here, we extend this curation to include the EPICv2 and MSA arrays. The resulting set of CpGs can provide a resource for future epigenetic clock development.

Second, we expanded the set of machine learning techniques used to model the clock. Our original Pace of Aging epigenetic clock was derived from a single elastic-net regression. Here, we applied an ensemble regularized regression approach, fitting 100 models per algorithm and averaging parameters to construct the final specification. This approach capitalizes on the observation that clocks distributing model weights across larger numbers of CpG sites have superior technical reliability^21^, a finding further confirmed in comparison of our Ridge and ElasticNet versions of the clock. The Ridge method retains information from all CpGs included in machine learning analysis, whereas the elastic net selects a subset. The superior technical reliability of the Ridge-model clock, combined with equal or superior healthspan prediction across datasets, suggests ridge regression may offer a promising approach for future clock development, at least in the context of curated CpG sets, such as the one used here. However, this increased technical reliability may come at the price of reduced sensitivity to intervention; the Ridge clock was less responsive to CALERIE intervention as compared with the ElasticNet model. This result could encourage use of the elastic-net model in shorter-term intervention studies. We provide access to both models through the FraminghamPACE Projector available through GitHub (https://github.com/will-marella/FraminghamPACE).

Third, we applied a statistical approach to exclude variation in calendar age from the clock parameters. Our original Pace of Aging epigenetic clock was developed within the Dunedin Study, in which all participants are the same calendar age. In the Framingham Heart Study Offspring cohort, participants’ ages vary by decades. As expected, older participants tended to have faster Pace of Aging. To ensure this correlation did not contaminate the epigenetic clock with CpGs associated with calendar age but not physiological change, we included participant age at DNAm collection as a covariate in machine learning analysis to select CpGs. This follows the method introduced in GrimAge^10^. However, in contrast to the GrimAge approach, we exclude the age parameter from our final algorithm, leaving only the age-independent CpG associations with our Pace of Aging phenotype. Strikingly, despite having excluded age variation from clock development, the resulting FraminghamPACE epigenetic clock was positively correlated with age and showed increases over time in within-participant analysis of change in the CARDIA cohort. Thus, even when statistically controlling age variation in clock development, the resulting Pace of Aging epigenetic clock remains sensitive to acceleration in biological aging across the adult lifespan.

We acknowledge limitations. The Framingham Offspring cohort is drawn from a single Massachusetts town and is almost entirely of white European ancestry. It does not represent the full US population. Validation analyses in diverse cohorts recruited from across the US, including CARDIA, HRS, and WHI, suggest that the resulting DNA methylation biomarker has significant generalizability. Nevertheless, further replication in cohorts from outside the US is needed, as are studies of younger populations. The biomarker panel available to measure Pace of Aging in Framingham, while extensive (17 biomarkers across vascular, inflammatory, renal, metabolic, pulmonary, hepatic, and hematologic systems), remains an incomplete summary of physiological changes occurring with aging. We excluded measurements of lipids and blood pressure because active medical management of these parameters was initiated during follow-up. Many measurements of the immune system were available only toward the end of follow-up. Other aspects of physiology such as bone and muscle function were not included. Expanded sets of measurements may improve future Pace of Aging analysis. DNA methylation data in the Framingham Offspring cohort were generated from an older technology (the Illumina Infinium 450k array). We focused our analysis on a set of CpG sites conserved across generations of Illumina technology and showed that the clock performed well in EPICv1 datasets from HRS, CARDIA, and CALERIE. Analysis of Pace of Aging in datasets with newer, higher-coverage arrays may yield superior biomarkers. Analysis of response to intervention was limited to the CALERIE trial. CALERIE is, to date, the only trial of long-term calorie restriction in healthy adults. Follow-up in new trials of interventions established to modify aging biology in laboratory models is needed.

Within the context of these limitations, FraminghamPACE demonstrates the feasibility and value of developing Pace of Aging speedometers from existing longitudinal cohorts. Together with DunedinPACE, it establishes Pace of Aging measurement as a valuable tool for intervention research and opens the door for developing additional speedometers in diverse populations to strengthen and extend this approach.

## METHODS

### Framingham Heart Study ODspring Cohort

We developed FraminghamPACE in the Framingham Heart Study Offspring Cohort. The Framingham Heart Study is a multi-generational cohort initiated in 1948^33^. We conducted Pace of Aging analysis in the Offspring Cohort (2^nd^ generation, n=5,124; baseline data collection beginning in 1971). Participants were on average 36 years of age at baseline and 48% were men. At this initial assessment and seven subsequent follow-up assessments, the study conducted physical examinations and collected blood samples. DNA from the eighth assessment was analyzed to generate the DNAm dataset (dbGaP Accession: phs000724.v9.p13; n=2,069).

### Validation Cohorts

We conducted follow-up analysis of FraminghamPACE in five additional studies. To evaluate the technical reliability of FraminghamPACE, we compared values in technical replicates from LOLIPOP (GSE55763, n=36 replicates) and CARDIA (dbGaP accession phs000285.v3.p2; n=38 replicates). To evaluate how FraminghamPACE varied across the lifespan, we analyzed repeated-measures DNAm data collected over 15y of follow-up in the CARDIA Study (dbGaP accession phs000285.v3.p2; n=2,023 participants with up to 4 time points of DNAm data; 6,214 DNAm observations in total). To compare performance of FraminghamPACE to DunedinPACE and other clocks in prediction of healthspan and lifespan, we analyzed data from the US Health and Retirement Study (HRS, NIAGADS Accession sa000021; N=3,483, 6 years of follow-up from DNAm collection) and the Women’s Health Initiative (WHI, dbGaP Accession phs000200.v12.p3; N=1,835, 25 years of follow-up from DNAm collection). Finally, to evaluate responsiveness to geroprotective intervention, we analyzed data from the CALERIE Randomized Controlled Trial^34^. Descriptions of individual cohorts are provided in the **Supplemental Methods**.

### DNAm Processing

DNAm data for CALERIE were processed at Duke University as described previously^35^. DNAm from the LOLIPOP study was obtained from the Gene Expression Omnibus (GSE55763). All other DNAm datasets were processed in the Columbia Aging Center Geroscience Computational Core in R (v 4.4.1) using the minfi (v 1.50.0)^36^ and ewastools packages (v 1.7.2)^37^. Details are reported in the **Supplemental Methods**.

### Calculated DNAm variables

We calculated FraminghamPACE in the validation datasets using the R package published on GitHub (https://github.com/will-marella/FraminghamPACE). We computed DNAm estimates of leukocyte fractions using the methods developed by Houseman^38^ and Salas^39^ via the R package published on GitHub (https://github.com/immunomethylomics/FlowSorted.BloodExtended.EPIC). We calculated DunedinPACE^11^ using the R package published on GitHub (https://github.com/danbelsky/DunedinPACE). We calculated PCGrimAge, PCPhenoAge, PCHorvath, and PCHannum using the PC Clocks R package published by Levine and colleagues^21^ (https://github.com/HigginsChenLab/methylCIPHER).

### Statistical Analysis

#### Pace of Aging

To measure Pace of Aging, we analyzed biomarker data for all participants contributing at least one measurement of any of the 17 biomarkers (n=4,988). We fit a hierarchical model to the biomarker data to estimate the Pace of Aging. In this approach, each biomarker (*k*) is modeled for individual (*i*) at timepoint (*t*) as: *y*_*itk*_ = *β*_0*ik*_ + *β*_1*ik*_*t* + *β*_2*ik*_*t*^2^ + *ϵ*_*itk*_ and each parameter *β* is estimated as *β*_*xik*_ = *μ*_*x*_ + *θ*_*xi*_ + *δ*_*xik*_ for *x* ∈ {0, 1, 2}. This model allows for both linear and quadratic slopes of change (*β*_1_, *β*_2_) and specifies a hierarchical structure consisting of cohort-level (*μ*), participant-level (*θ*), and participant-biomarker-level (*δ*) components. We extracted the participant-level linear slope shared across all biomarkers (*θ*_1*i*_, i.e. the *θ* corresponding to *β*_1_, + the population-level linear slope, *μ*_1_) for the n=2,705 Offspring Cohort members with at least two measurements of 13 or more of the 17 biomarkers. We scaled this parameter separately by sex to reflect years of biological change experienced by the participant per calendar year. To do this, we first divided each participant’s value by the sex-specific cohort mean^8^. We next scaled the resulting ratios by the survival function for age in the cohort so that the implied years of biological aging over the 30-year follow-up (i.e. 30 for Pace of Aging=1, 60 for Pace of Aging=2) corresponded to the mortality hazard for the equivalent change in calendar age. Specifically, we fitted Gompertz models of survival for the ratio metric (*μ*(*t*)_*tatio*_) and separately for chronological age (*μ*(*t*)_*age*_). We scaled Pace of Aging as 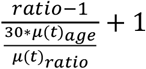. The scaled Pace of Aging value retains a mean of 1, but the variance is rescaled such that units correspond to the mortality risk implied by 30 years of aging. Details of participants included in each phase of Pace of Aging analysis are reported in **Supplemental Table 2**.

We modeled FraminghamPACE using DNA methylation data from whole blood collected at the Framingham Heart Study Offspring Cohort’s eighth assessment wave. CpG sites were first filtered for cross-platform technical reliability^32^, yielding 14,465 CpGs used for model development (**Supplemental Methods**; **Supplemental Table 4)**.

#### FraminghamPACE

We modeled FraminghamPACE in the subset of FHS participants with a Pace of Aging phenotype and DNAm data (n = 2,069). We fit elastic-net and ridge regression models using the ‘glmnet’ R package^40^. We included age and sex as unpenalized covariates.

##### Ensemble Machine Learning

Our ensemble machine learning approach followed the “bagging” approach^20^. We first selected the regularization hyperparameter for each model using 10-fold cross-validated grid search. We next fit 100 iterations of each model. These iterations utilized the selected hyperparameter and were each fit to an independent random 90% subsample of the training data. (In the original implementation of bagging, iterations are performed on bootstrapped samples, which can include the same participant multiple times within a given iteration.) Finally, we averaged coefficients across models. In the ridge models, every CpG has a non-zero coefficient in every model. In the elastic-net model, only a subset of CpGs have a non-zero coefficient in each model. When calculating ensemble parameters for the elastic net model, we averaged across non-zero and zero coefficients to compute the final parameter. This approach stabilizes coefficient estimates, improving model fit, robustness, and generalizability^41^. Training results are summarized in **Supplemental Table 5**. The final FraminghamPACE algorithms omit age and sex parameters, yielding age- and sex-independent DNAm scores (https://github.com/will-marella/FraminghamPACE).

##### Reliability and Validation Analysis

We conducted analysis of technical reliability by calculating intraclass correlation coefficients (ICCs) for replicate DNAm samples using the ‘rptR’ R package^42^. We conducted analysis of variation across the adult lifespan using ordinary least squares regression in cross-sectional data and fixed-effects (or “within-subjects”) regression in longitudinal repeated measures data using the ‘plm’ R package^43^ and the ‘lmtest’ R package^44^. We conducted analysis of healthspan and lifespan using Cox proportional hazards models for time-to-event outcome data, Poisson regression models for count outcome data, and multinomial logistic regression for categorical outcome data using the base R ‘stats’ package^45^, ‘survival’ R package^46^, and ‘nnet’ R package^47^. We conducted analysis of response to intervention by fitting mixed models via the ‘lme4’ R package^48^. Additional details are noted in the **Supplemental Methods**.

## Data Availability

Data for the Framingham Heart Study (FHS) are available from dbGaP (Accession: phs000724.v9.p13; we analyzed the version 33 release of these data).
Data from LOLIPOP are available from the Gene Expression Omnibus (GEO Accession: GSE55763).
Data from the Coronary Artery Risk Development in Young Adults (CARDIA) are available from NHLBI BioLINCC (Accession HLB00030026a) and dbGaP (Accession phs000285.v3.p2).
Data from the US Health and Retirement Study (HRS) are available from the University of Michigan Institute for Social Research (https://hrs.isr.umich.edu/data-products), NIAGADS (https://dss.niagads.org/datasets/ng00153/; Accession sa000021), and the NIH database of Genotypes and Phenotypes (dbGaP; Accession phs000428.v2.p2).
Data from the Women's Health Initiative (WHI) are available from dbGaP (Accession: phs000200.v12.p3).
Data from the CALERIE trial are available from the Aging Research Biobank (https://agingresearchbiobank.nia.nih.gov/studies/calerie) and the CALERIE Biorepository (https://calerie.duke.edu).

## Data Sharing Statement

Data for the Framingham Heart Study (FHS) are available from dbGaP (Accession: phs000724.v9.p13; we analyzed the version 33 release of these data).

Data from LOLIPOP are available from the Gene Expression Omnibus (GEO Accession: GSE55763).

Data from the Coronary Artery Risk Development in Young Adults (CARDIA) are available from NHLBI BioLINCC (Accession HLB00030026a) and dbGaP (Accession phs000285.v3.p2).

Data from the US Health and Retirement Study (HRS) are available from the University of Michigan Institute for Social Research (https://hrs.isr.umich.edu/data-products), NIAGADS (https://dss.niagads.org/datasets/ng00153/; Accession sa000021), and the NIH database of Genotypes and Phenotypes (dbGaP; Accession phs000428.v2.p2).

Data from the Women’s Health Initiative (WHI) are available from dbGaP (Accession: phs000200.v12.p3).

Data from the CALERIE trial are available from the Aging Research Biobank (https://agingresearchbiobank.nia.nih.gov/studies/calerie) and the CALERIE Biorepository (https://calerie.duke.edu).

